# Knowledge awareness and practice with antimicrobial stewardship programmes among healthcare providers in a Ghanaian Tertiary Hospital

**DOI:** 10.1101/2021.11.14.21266285

**Authors:** Eneyi E. Kpokiri, Misha Ladva, Cornelius C. Dodoo, Emmanuel Orman, Thelma Alalbila, Adelaide Mensah, Jonathan Jato, Kwadwo A. Mfoafo, Isaac Folitse, Araba Hutton-Nyameaye, Inemesit Okon-Ben, Paapa Mensah-Kane, Emmanuel Sarkodie, Benedict Awadzi, Yogini Jani

## Abstract

Antimicrobial resistance (AMR) is a significant problem in global health today, particularly in Low- and Middle-Income Countries (LMICs) where antimicrobial stewardship programmes are yet to be successfully implemented. We established a partnership between AMR pharmacists leads from a UK NHS hospital and in Ho Teaching Hospital with the aim to by enhance antimicrobial stewardship knowledge and practice among healthcare providers through an educational intervention. We employed a mixed method approach including an initial before and after training survey on knowledge and awareness, followed by qualitative interviews with healthcare providers conducted six months after delivery of training. This study was carried out in Ho teaching hospital with 18 healthcare professionals including pharmacists, medical doctors, nurses and medical laboratory scientists. Ethical approval was obtained prior to data collection. In the first phase, we surveyed 50 health care providers including nurses (33%), pharmacists (29%) and biomedical scientists (23%). Of these, 58% of participants had engaged in continuous professional development on AMR/AMS, and above 95% demonstrated good knowledge on the general use of antibiotics. A total of 18 participants, which included 4 medical doctors, 5 pharmacists, 4 nurses, 2 midwives and 3 biomedical scientists, were interviewed in the second stage and demonstrated greater awareness of AMS practices, particularly the role of education for patients as well as healthcare professionals. We found that knowledge and practice with AMS was markedly improved six months after the training session. There is limited practice of AMS in LMICs, however through AMR focused training we demonstrate improved AMS skills and practice among health care providers in Ho Teaching hospital. There is need for continuous AMR training sessions for health care professionals in resource limited settings.

## 1. Introduction

The introduction should briefly place the study in a broad context and highlight why it Antimicrobial resistance (AMR) is a significant problem in global health today. It is severe in Low- and Middle-Income Countries (LMIC’s) where the burden of infectious diseases is much higher. There is limited information on the extent of this problem in these settings. Global mortality projections due to AMR in the next few decades show greatest impact will be in Africa and Asia.^1^ Inappropriate prescribing and use of antibiotics have been shown to contribute significantly to this problem. Antimicrobial stewardship programmes (AMS) are designed to improve use of antimicrobials and they have been developed and implemented successfully with corresponding impact in most high-income settings. In 2015, the WHO released a Global Action Plan with a combination of AMS interventions which is yet to be fully implemented and tested in most LMICs due to lack of required resources.^2^ There is an urgent need to identify practical strategies to support delivery of AMS in these settings.

Widespread AMR has been documented in Ghana in national reports and published research.^3^ Ghana’s first national action plan for AMR to span from 2017 to 2021 was published in 2017 by the Ghanaian ministry of health with the support of WHO and the Food and Agriculture Organization of the United Nations. Multiple drug resistance is as high as 78% and resistance to common antimicrobials like tetracycline, cotrimoxazole and ampicillin is around 70%. High prevalence of MRSA has also been documented.^3^ Ghana’s national action plan outlines strategies for implementing stewardship to include monitoring and surveillance, education and training and provision of guidelines.^4^ Reports from Ghana have highlighted gaps in optimal antimicrobial use even in hospital.^5^ There is need to optimise antibiotic prescribing practices and strengthen monitoring, surveillance and documentation of antibiotics use and related processes. There is a paucity of evidence on the knowledge and extent to which healthcare staff in Ghana are involved with antimicrobial stewardship programmes.

The Tropical Health and Education Trust (THET) together with the Commonwealth Partnerships for Antimicrobial Stewardship (CwPAMS) developed and funded the partnership model between UK NHS hospitals and African partners.^6^ The goal of the partnerships was to leverage the NHS expertise to address AMR challenges with the input and resources from the African partners. This paper reports part of the activities and results of one of the partnerships between the University London College Hospital NHS foundation trust (UCLH) and the University of Health and Allied Sciences (UHAS) and Ho Teaching hospital, both in Ho municipality in the Volta region of Ghana. The activities carried in this partnership included periodic Global Point Prevalence Survey (GPPS)^7^ to establish patterns of antimicrobial use and an AMR/AMS training and capacity building workshop for healthcare professionals. The aim of this study was to improve AMS knowledge and build the capacity and skills of healthcare professionals to support the implementation of antimicrobial stewardship programmes in Ho Teaching Hospital in the Volta region of Ghana

## 2. Methods

### 2.1 Study setting

This study was conducted in Ho, teaching hospital located in the Volta region of Ghana. The hospital is a tertiary care hospital, serves as a teaching hospital with a staff strength of about 1200, 306 bed spaces and 14 wards. Ghana is low- and middle-income country located in sub-Saharan part of west Africa. Ghana has an established National Health Insurance Scheme (NHIS) which provides a broad range of health care services to Ghanaians through district mutual and private health insurance schemes.^8^

### 2.2 Study design

We employed a bottom-up approach mixed-methods study design. This included an initial survey of healthcare providers KAP on antimicrobial resistance and AMS, just before and after the delivery of an educational intervention. This was then followed with semi-structured interviews six months after the training to assess changes in AMR/AMS knowledge, awareness and improvements to practice. Partners from UCLH with AMR practice and research expertise developed and delivered AMR/AMS and quality improvement focused trainings over a 3-day period.

### 2.3 Data collection

For phase 1, we designed a data collection tool (supplemental file 1) to retrieve data on knowledge, awareness and practice of AMR/AMS among health care providers. For phase 2, an interview topic guide was designed (supplemental file 2) and used to administer the interviews. Prior to interviews, participants were provided an invitation and information leaflet with relevant information about the study as an invitation to participate. Participants were also provided a consent form to sign, and all interviews were audio recorded.

### 2.4 Data analysis

Data collected from the initial before and after training survey were entered into and analysed using the IBM Statistical Package for Social Sciences (SPSS) version 22. Results were presented using basic descriptive frequencies. All interviews were transcribed verbatim, and transcripts were read repeatedly and then coded individually to identify similarity and differences. All transcripts were coded and reviewed alongside coding frame generated. Codes were also mapped unto the Behaviour Change Wheel (BCW) framework based on the influence of Capability, Opportunity and Motivation on Behavioural outcomes (COM-B) model (see Figure 1 below).^9^ Similar codes were grouped for themes and high-level findings which were presented as a narrative using quotes to support the main themes. To ensure validity, all transcripts were coded by the researcher and randomly validated by another team member.

**Figure 1:**
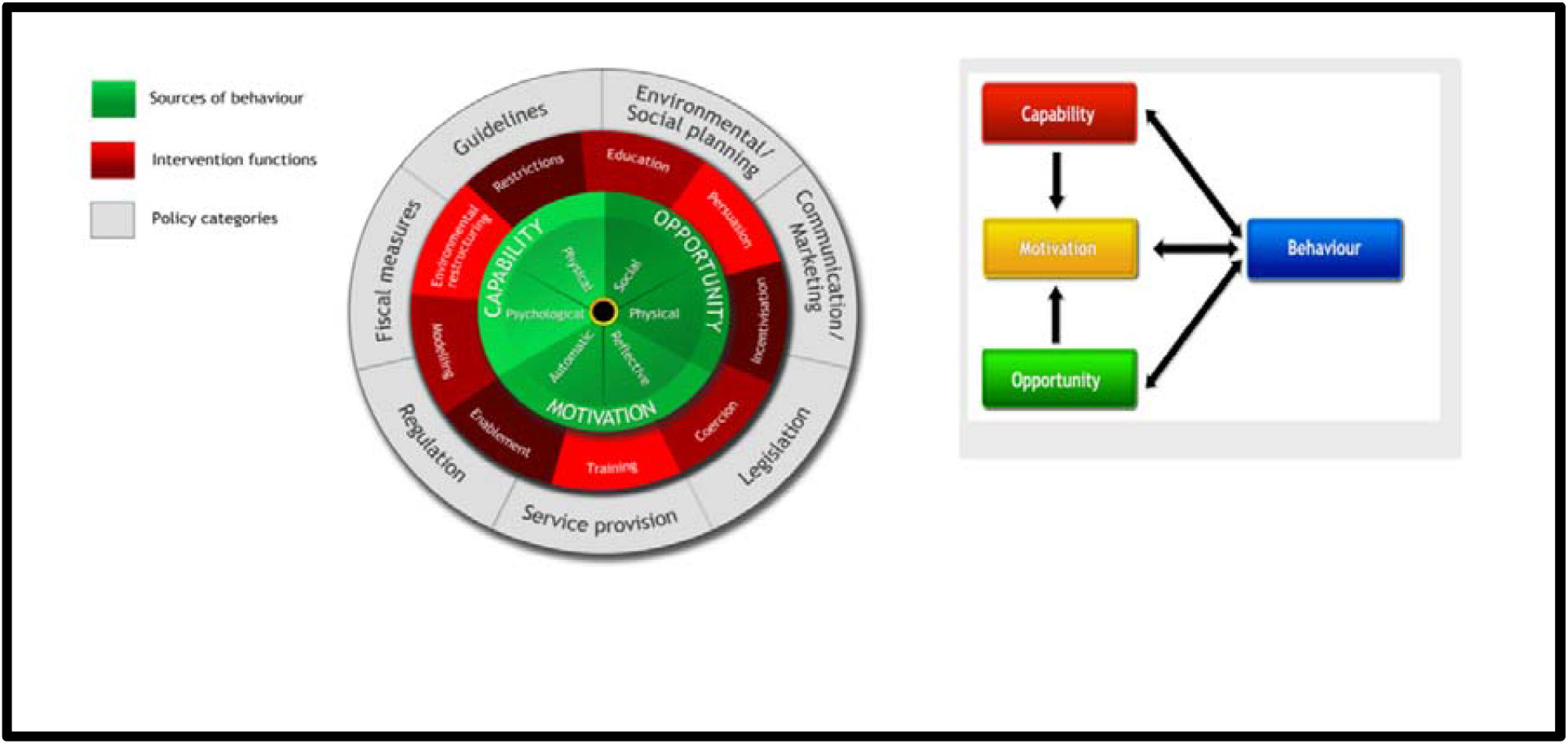
The Behaviour Change Wheel and COM-B model.

### 2.5 Ethical approval

Ethical approval was obtained via ethics application made to the research and ethics committee of the University of Health and Allied Sciences.

## 3. Results

*Phase 1: AMR knowledge awareness and practice pre and post an AMR/AMS training session*

### 3.1 Socio-demographics

A total of 50 healthcare professionals attended the AMR training session including nurses (33%), Pharmacists (29%) and medical laboratory scientists (23%). Of these, 96% were aged between 25-44years with slightly more females (54%) than males (46%). Most participants (96%) have achieved up to master’s degree level education.

**Table 1:** Socio-demographics of participants.

| **Characteristics (n = 50)** | **N (%)** |
| --- | --- |
| **Gender** |  |
| Male | 23 (46) |
| Female | 27 (55) |
| **Age (years)** |  |
| 25-34 | 33 (66) |
| 35-44 | 15 (30) |
| 45-54 | 1 (2.0) |
| 55-64 | 1 (2.0) |
| **Occupation** |  |
| Nurse | 16 (33.3) |
| Pharmacist | 14 (29.2) |
| Biomedical Scientist | 11 (22.9) |
| Medical doctor | 3 (6.3) |
| Pharmacy Technologist | 1 (2.08) |
| Midwife | 1 (2.08) |
| Lecturer | 1 (2.08) |
| Human Resource Manager | 1 (2.08) |

### 3.2 AMR Knowledge and awareness

Up to 58% of participants engage in CPD with topics on AMR/AMS, and above 95% demonstrated good knowledge on the general use of antibiotics. While many were familiar with terms such as antibiotic resistance and antimicrobial resistance, more than half were not conversant with antimicrobial stewardship, super bug and AMR capacity building.

**Table 2:** Familiarity with AMR/AMR terminology.

| **Indicator** | **Yes** | **No** | **Not Applicable** |
| --- | --- | --- | --- |
| *Antibiotic resistance* | 50 | 0 | 0 |
| *Antimicrobial capacity building* | 1 | 49 | 0 |
| *Antimicrobial resistance* | 45 | 3 | 0 |
| *Antimicrobial stewardship* | 22 | 12 | 8 |
| *Superbugs* | 14 | 19 | 7 |
| *Drug resistance* | 49 | 1 | 0 |

### 3.3 AMR Practice

More than 80% of participants agreed that patients’ clinical conditions and microbiology results were key determinants for antibiotic prescriptions, while more than 97% agreed there were lack of effective diagnostic tools and that inappropriate prescribing with antibiotics still exists. About half of the participants confirmed that there are no specific prescribing policies and protocols and antibiotics available in the facility are unable to treat some infections. The national standard treatment guideline (STG) is the most consulted reference when prescribing antibiotics followed by the British National Formulary (BNF). Over 90% of participants agreed that obtaining local antibiotic resistance profiles and changing the attitudes of prescribers and patients will reduce unnecessary antibiotic usage.

*Phase 2: Interviews with healthcare providers in HTH*

#### Participant characteristics

For the second phase, we interviewed a total of 18 participants, which included 4 medical doctors, 5 pharmacists, 4 nurses, 2 midwives and 3 biomedical scientists. Among the participants were 10 males and 8 females with age ranging from 27 to 56years.

#### Knowledge/Awareness of antimicrobial resistance post training

Majority of the interview participants confirmed that the training on AMR/AMS was very helpful and increased their understanding of AMR. This has in turn led to changes in their practice such as reduced empirical prescribing of antibiotics and more detailed counselling for patients on antibiotics use.

#### Healthcare provider roles

The healthcare providers across different specialties explained that they feel more confident carrying out AMS roles after the training. Nurses are competent in antibiotics drug administration and act as patient advocate to question antibiotics being prescribed. The pharmacists are also competent to deliver AMS roles and provide medication information needed to rational antibiotic prescribing. Both nurses and pharmacists can support antibiotic prescribing decisions especially when it involves the junior doctors.

#### Current situation with antibiotics use in Ghana

Interview participants reported general overuse of antibiotics in Ghana with high empirical and broad-spectrum prescribing. Factors that contribute to this observed practice with antibiotics include lack of continuous training for prescribers and other HCPs, poor compliance with guidelines, limited diagnostic services and staff shortages, pressure from patients and influence of drug companies. Till date there has been no interventions put in place to improve antibiotics use in the study hospital.

#### Prescribing patterns and the decision-making process

Commonly prescribed antibiotics include the Cephalosporins, penicillin’s, metronidazole, ciprofloxacin, erythromycin, amoxiclav and cefazolin. All of these antibiotics are in the essential medicines list and available in hospital formulary both as branded and generic forms. The common indications requiring antibiotic prescriptions include UTI, respiratory tract infections, GIT infections, sepsis, STIs and surgical prophylaxis. There are standard treatment guidelines, however, the disease type, cost, drug availability and severity of infection are all factors that affect the choice of antibiotic drug being prescribed.

#### Recommended strategies to improve antibiotics use and the barriers in implementing these AMS interventions

Respondents highlighted some recommendations to improve antibiotics use as including education and training for prescribers, Improved labs for microbe specific treatment, purchase lab items for testing, antibiotics use checks/audits, employment of more staff to build workforce and development of local policies. Main barriers to this will be failure to enforce laws and limited funding in healthcare.

### Mapping Codes and themes to the BCW and COM-B Model

| Mapping Codes and themes to the BCW and COM-B Model |  |  |  |
| --- | --- | --- | --- |
| Themes | BCW framework |  | Sources of Behaviour (COM-B) |
|  | Intervention Functions | Policy Category |  |
| Factors contributing to irrational use of antibiotics include |  |  |  |
| Lack of continuous AMR trainings | Training | Social planning | Psychological Capability |
| Poor AMR knowledge | Enablement |  | Psychological Capability |
| Limited diagnostic services |  | Service provision | Physical capabilities |
| Staff shortages |  | Service provision |  |
| Pressure from patients | Persuasion/Coercion |  | Reflective motivation |
| Incentives from Pharmaceutical companies | Incentivisation | Communication marketing |  |
| Recommended strategies to improve antibiotics use |  |  |  |
| Education to increase awareness | Education/Enablement |  | Automatic motivation |
| Checks/audits / Monitoring antibiotic use |  | Regulations |  |
| Upgrade laboratory services /Purchase lab items for testing | Environmental restructuring |  | Physical Opportunity |
| Policies | Enablement | Guidelines | Automatic motivation |
| Training for prescribers | Training /Enablement |  | Automatic motivation |
| Employ more staff to build workforce, | Environmental restructuring | Social planning/ Service provision | Physical Opportunity |
| Barriers in implementing these AMS intervention |  |  |  |
| Lack of funding in healthcare |  | Fiscal measures |  |
| Staff shortages |  |  |  |
| Failure to enforce laws |  | Regulation/Legislation |  |

#### Sources of Behaviour

From the interview data, the current AMR behaviours we see such has high broad spectrum and empirical prescribing of antibiotics fall on the inherent local factors include lack of continuous training, limited laboratory services and lack of local policies. In other to move towards the desired behaviours for AMS support and practice we need to leverage the relevant intervention functions behaviour on the wheel. These include training, enablement, persuasion, and Environmental restructuring. These functions map unto the Social planning / Service provision, guidelines and regulations policy categories.

##### Capability

HCPs feel more confident and empowered to contribute on AMR topics during rounds and educate patients better after the training

##### Opportunity

Employ more staff to create opportunities to deliver additional AMS activities

##### Motivation

Training was educative, increased knowledge about antibiotics use and helped to improve AMS practice

## 4. Discussion

HCPs described the training as educative, helpful and found it useful in their practice with antibiotics. The training generally increased knowledge about antibiotics use, AMR and AMS. Led to better understanding of different aspects of antibiotics use including importance of proper patient education. After the training, some changes in practice observed in the hospital include reduction in antibiotics use and empirical prescribing. Better patient counselling and education on antibiotics use. HCPs feel more confident to contribute on AMR topics during rounds and educate patients better. Pharmacists carry out prescription checks for antibiotics prescribed and query antibiotics prescribed where there is a need too. The GPPS conducted made prescribers more aware of their prescribing with regards to antibiotics. There was reduce empirical and broad-spectrum antibiotic prescribing. However, majority of the HCP that attended the earlier training vaguely remembered the WHO classification of antibiotics

Pharmacists are the custodians of drugs within the hospital and can ensure safe effective use of antibiotics by providing information to other HCPs, counselling patients. Pharmacists are also competent to deliver AMS roles and provide medication information needed to rational Abx prescribing. Nurses are competent in antibiotics drug administration and act as patient advocate to question antibiotics being prescribed. While there are guidelines and consultation with other colleagues, HCPs confirmed other resources like improved laboratory services, and increase in manpower will increase efficiency in delivery of AMS roles. HCPs recommended protocols and guidelines for ABX use, more AMR training courses, improved lab services and availability of patient educational materials to support the delivery of AMS in the local hospital.

HCPs were of the perceptions that there is a general there is irrational and overuse of antibiotics in Ghanaian hospitals citing broad spectrum and empirical prescribing of antibiotics. Factors contributing to irrational use of antibiotics in Ghana include lack of continuous AMR trainings, poor AMR knowledge, limited diagnostic services, understaffing, pressure from patients and pharmaceutical companies.

Broad-spectrum antibiotics including penicillin’s, cephalosporins aminoglycosides and metronidazole are commonly prescribed. Common indications for which antibiotics are prescribed include respiratory tract infections, UTI, STI, GIT infections. Others include typhoid and surgical prophylaxis. In the study hospital, antibiotics are prescribed both as the brands and in generics but mostly in generics and are in the EML and hospital formulary.

Presently, there are standard treatment guidelines in the hospital to guide antibiotics use. Most HCP employ the use of guidelines as often as they need to when dispensing, prescribing or administering antibiotics. While patients can request for antibiotics, they have no influence in the decision whether they eventually get an antibiotic or which antibiotic should be prescribed for and dispensed to them. In addition to laboratory diagnostics reports, the following factors are considered in making choice of antibiotics for a patient: the disease and its severity, cost of the antibiotic and availability.

The findings of this work have several implications for practice and policies. Given the factors underpinning current antimicrobial use patterns, there is need for dedicated AMS staff to promote and oversee AMS activities in the hospital. Routine trainings and AMR focused workshops should be organised for healthcare providers to reinforce and sustain AMS practices. Following data from GPPS results, local AMS policies should be developed to guide antimicrobial use in the local hospital context. National and international treatment guidelines should be adapted and tailored for relevance and use in the local settings. There is need for further research to evaluate the impact of the AMS team on practice and to identify further parameters for long term sustainability.

Strengths and limitations

## Conclusion

AMR is still a budding public health challenge across the globe and especially in low-income settings. AMS practice is still in infancy in these settings. We demonstrate how expertise from developed and advance settings can be leveraged to move AMS practice forwards. Findings from this study shows continuous AMR training for healthcare professionals can increase knowledge and awareness, spur behaviour change to improve practice and build momentum for sustainable AMS programmes.

## Supporting information

Supplemental foles

## Data Availability

All data produced in the present study are available upon reasonable request to the authors

## Supplemental materials

Appendix I: Survey questionnaire for antibiotic use and antimicrobial resistance

Appendix II: Healthcare providers’ interview topic guide

Appendix III: AMS Interview data analysis

Appendix IV: Sample transcript Coding frame

## Author Contributions

Conceptualization, EEK, YJ, AM, KAM, EO, AH, JJ, CCD, TA, ES, IF, AHN, IOB, PMK, ML. ; methodology: EEK, YJ and ML; Survey development: CCD, EO, EEK, TA, AM, IF, KAM, BA formal analysis; EEK, ML, BA, EO, TA, JJ, IF, AH, IO, PM, ES, ML, YJ; data curation, CCD, EO, TA, AM, IF, KAM, BA, YJ; writing—original draft preparation, EEK ; writing—review and editing, EEK, YJ, AM, EO, AH, JJ, CCD, TA, ES, IF ; project administration, CCD, EEK, JJ, AM, EO, YJ; funding acquisition, EEK, CCD and YJ. All authors read and approved the final version of the manuscript.

## Funding

This project was funded by the Commonwealth Partnerships for Antimicrobial Stewardship (CwPAMS) through the Tropical Health and Education Trust (THET) and the Fleming fund. Grant number A07 (2019)

## Acknowledgments

The authors would like to thank the staff of Ho Teaching Hospital Pharmacy department and the Faculty of Pharmacy at the University of Health and Allied Sciences, Ho for their support throughout this work.

## Conflict of interest

The authors declare no conflicting interests

## Notes

### Competing Interest Statement

The authors have declared no competing interest.

