## Supplemental foles for "Knowledge awareness and practice with antimicrobial stewardship programmes among healthcare providers in a Ghanaian Tertiary Hospital"

**Appendix I**

**SURVEY QUESTIONNAIRE FOR ANTIBIOTIC USE AND ANTIMICROBIAL RESISTANCE**

This form has been designed and validated for the use to collect data from health professionals on their knowledge and effective use of antimicrobials for the purposes of training as part of an ongoing project on Antimicrobial Stewardship. We are particularly interested in knowing more about antibiotic use and antibiotic resistance in your health facility; your knowledge and how you manage it. We are also interested in any problems or concerns you might have about antibiotics use. We would appreciate it if you could spend about 10 minutes with us discussing the topic.

1. **BACKGROUND INFORMATION ON THE HEALTH PROFESSIONAL**

*Kindly tick against the appropriate option applicable to you.*

| **A** | Sex | Male 󠆿 | | | 🞎 | Female | | | 🞎 | |  | |  |
| --- | --- | --- | --- | --- | --- | --- | --- | --- | --- | --- | --- | --- | --- |
| **B** | Age (Years) | 19 to 24 | | | 🞎 | 25 to 34 | | | 🞎 | | 35 to 44 | | 🞎 |
|  |  | 45 to 54 | | | 🞎 | 55 to 64 | | | 🞎 | | >64 | | 🞎 |
| **C** | Highest degree or level of school completed | No schooling completed | | | 🞎 | Junior high school level | | | 🞎 | | Senior high school | | 🞎 |
|  |  | Technical / vocational school | | | 🞎 | Tertiary level, diploma or HND degree | | | 🞎 | | Tertiary level, bachelor degree | | 🞎 |
|  |  | Tertiary level, master’s or professional degree | | | 🞎 | Tertiary level, doctorate degree | | | 🞎 | | Other  specify: _________ | | 🞎 |
| **D** | Profession | Medical Doctor | | | 🞎 | Physician Assistant | | | 🞎 | | Nurse | | 🞎 |
|  |  | Pharmacist | | | 🞎 | Pharmacy Technologist | | | 🞎 | | Other  specify: _________ | | 🞎 |
| **E** | Years of practice | < 1 year | 🞎 | 1 to 5 years | | | 🞎 | 6 to 10 years | | 🞎 | | > 10 years | 🞎 |
| **F** | Number of years working at the present facility | < 1 year | 🞎 | 1 to 5 years | | | 🞎 | 6 to 10 years | | 🞎 | | > 10 years | 🞎 |

2. **HOSPITAL PRACTICE REGARDING THE USE OF ANTIMICROBIALS**

*Kindly tick against the appropriate option applicable to you*

| **Sr. No** | **Information** | **Strongly Agree** | **Agree** | **Not sure** | **Disagree** | **Strongly Disagree** |
| --- | --- | --- | --- | --- | --- | --- |
| **A** | The following factors influence the decision to start a patient on antimicrobial therapy in the hospital |  |  |  |  |  |
|  | i. *Patient’s clinical condition* | 🞎 | 🞎 | 🞎 | 🞎 | 🞎 |
|  | ii. *Microbiological results in symptomatic patients* | 🞎 | 🞎 | 🞎 | 🞎 | 🞎 |
| **B** | The following practices contribute to antimicrobial resistance in the hospitals |  |  |  |  |  |
|  | i. *Inappropriate prescribing habits of antibiotics* | 🞎 | 🞎 | 🞎 | 🞎 | 🞎 |
|  | ii. *Lack of effective diagnostics tools to diagnose bacterial infections* | 🞎 | 🞎 | 🞎 | 🞎 | 🞎 |
|  | iii. *Patients self-medication with antibiotics without consulting health professionals* | 🞎 | 🞎 | 🞎 | 🞎 | 🞎 |
|  | iv. *Spread of bacteria in healthcare settings due to poor hygiene practices* | 🞎 | 🞎 | 🞎 | 🞎 | 🞎 |
| **C** | Antibiotics are overprescribed in this facility | 🞎 | 🞎 | 🞎 | 🞎 | 🞎 |
| **D** | Antibiotics choice should only be made base on laboratory results always | 🞎 | 🞎 | 🞎 | 🞎 | 🞎 |
| **E** | Current antibiotics available in the facility are unable to treat some infections | 🞎 | 🞎 | 🞎 | 🞎 | 🞎 |
| **F** | There are policies and protocols for antibiotic use in this facility | 🞎 | 🞎 | 🞎 | 🞎 | 🞎 |
| **G** | All prescriptions are based on the hospitals protocol | 🞎 | 🞎 | 🞎 | 🞎 | 🞎 |
| **H** | Poor infection control practices by healthcare professionals will cause the spread of antimicrobial resistance. | 🞎 | 🞎 | 🞎 | 🞎 | 🞎 |
| **I** | The following practices may help control antimicrobial resistance |  |  |  |  |  |
|  | i. *Consulting with infectious diseases experts* | 🞎 | 🞎 | 🞎 | 🞎 | 🞎 |
|  | ii. *Obtaining local antibiotic resistance profile* | 🞎 | 🞎 | 🞎 | 🞎 | 🞎 |
|  | iii. *Targeting antimicrobial therapy to likely pathogens* | 🞎 | 🞎 | 🞎 | 🞎 | 🞎 |
|  | iv. *Changing the attitudes of prescribers and patients to reduce unnecessary antibiotic usage* | 🞎 | 🞎 | 🞎 | 🞎 | 🞎 |

| **J** | In using antibiotics in the hospital, the following resources are consulted (enter 1, 2, or 3 based on the order in which you would consult these) | | | | |
| --- | --- | --- | --- | --- | --- |
|  | Google | 🞎 | Standard Treatment Guidelines | 🞎 |  |
|  | Colleagues | 🞎 | British National Formulary | 🞎 | Other 🞎 Specify  ______________________________ |

**K**	Is there an antimicrobial stewardship (AMS) team in your hospital? **Yes** 🞎	No 🞎

If yes, Please state the following

I. Composition of the team

| Medical Doctor | 🞎 | Physician Assistant | 🞎 | Nurse | 🞎 |
| --- | --- | --- | --- | --- | --- |
| Pharmacist | 🞎 | Pharmacy Technologist | 🞎 | Other  specify: _________ | 🞎 |

II. Frequency of meetings	______________________________
III. Are minutes taking during meetings? **Yes** 🞎	No 🞎
IV. List the activities undertaken by the AMS team ______________________________
3. **GENERAL KNOWLEDGE ABOUT ANTIBIOTICS AND THEIR USE**

*Kindly tick against the appropriate option applicable to you*

| **Sr No** | **Information** | **Strongly Agree** | **Agree** | **Not sure** | **Disagree** | **Strongly Disagree** |
| --- | --- | --- | --- | --- | --- | --- |
| **A** | Antibiotics are used in the management of all infections | 🞎 | 🞎 | 🞎 | 🞎 | 🞎 |
| **B** | Treatment with antibiotics should be stopped once you feel better, especially the expensive ones | 🞎 | 🞎 | 🞎 | 🞎 | 🞎 |
| **C** | It’s okay to use antibiotics that were given to a friend or family member, as long as they were used to treat the same illness | 🞎 | 🞎 | 🞎 | 🞎 | 🞎 |
| **D** | It’s okay to buy the same antibiotics, or request these from a doctor, if you’re sick and they helped you get better when you had the same symptoms before” | 🞎 | 🞎 | 🞎 | 🞎 | 🞎 |
| **E** | Frequent use of antibiotics may decrease the efficacy of treatment | 🞎 | 🞎 | 🞎 | 🞎 | 🞎 |
| **F** | Antibiotics use should be strictly controlled | 🞎 | 🞎 | 🞎 | 🞎 | 🞎 |
| **G** | The following factors may influence the inappropriate use of antibiotics |  |  |  |  |  |
|  | i. *Counselling of patients* | 🞎 | 🞎 | 🞎 | 🞎 | 🞎 |
|  | ii. *Skills and knowledge of prescribers* | 🞎 | 🞎 | 🞎 | 🞎 | 🞎 |
|  | iii. *Patient self-medication* | 🞎 | 🞎 | 🞎 | 🞎 | 🞎 |
|  | iv. *Inadequate supervision with the medicine administration* | 🞎 | 🞎 | 🞎 | 🞎 | 🞎 |
| **H** | It is possible for the antibiotics we are using today to stop working properly in the future | 🞎 | 🞎 | 🞎 | 🞎 | 🞎 |

**I**. The following conditions can be treated with antibiotics (*you may tick more than one, if appropriate*)

| HIV/AIDS | 🞎 | Bladder infection or urinary tract infection (UTI) | 🞎 | Cold and flu | 🞎 |
| --- | --- | --- | --- | --- | --- |
| Gonorrhoea | 🞎 | Diarrhoea | 🞎 | Fever | 🞎 |
| Malaria | 🞎 | Measles | 🞎 | Skin or wound infection | 🞎 |
| Sore throat | 🞎 | Body aches | 🞎 | Headaches | 🞎 |

J. What does the WHO AWaRE categories for antibiotics stand for?
4. **AWARENESS OF ANTIMICROBIAL RESISTANCE and STEWARDSHIP**

*Kindly tick against the appropriate option applicable to you*

| **Sr. No** | **Information** | **Strongly Agree** | **Agree** | **Not sure** | **Disagree** | **Strongly Disagree** |
| --- | --- | --- | --- | --- | --- | --- |
| **B** | Antibiotic resistance occurs when your body becomes resistant to antibiotics and they no longer work as well | 🞎 | 🞎 | 🞎 | 🞎 | 🞎 |
| **C** | Many infections are becoming increasingly resistant to treatment by antibiotics | 🞎 | 🞎 | 🞎 | 🞎 | 🞎 |
| **D** | If bacteria are resistant to antibiotics, it can be very difficult or impossible to treat the infections they cause | 🞎 | 🞎 | 🞎 | 🞎 | 🞎 |
| **E** | Antibiotic resistance is an issue that could affect me or my family | 🞎 | 🞎 | 🞎 | 🞎 | 🞎 |
| **F** | Antibiotic resistance is an issue in other countries but not here | 🞎 | 🞎 | 🞎 | 🞎 | 🞎 |
| **G** | Antibiotic resistance is only a problem for people who take antibiotics regularly | 🞎 | 🞎 | 🞎 | 🞎 | 🞎 |
| **H** | Bacteria which are resistant to antibiotics can be spread from person to person | 🞎 | 🞎 | 🞎 | 🞎 | 🞎 |
| **I** | Antibiotic-resistant infections could make medical procedures like surgery, organ transplants and cancer treatment much more dangerous | 🞎 | 🞎 | 🞎 | 🞎 | 🞎 |
| **J** | Inappropriate use of antibiotics can lead to antibiotic resistance | 🞎 | 🞎 | 🞎 | 🞎 | 🞎 |
| **K** | Inappropriate use of antibiotics can lead to increased adverse effects and additional burden | 🞎 | 🞎 | 🞎 | 🞎 | 🞎 |

**A**	List three to five barriers to antimicrobial stewardship and suggest ways to overcome these.

(In particular, please highlight barriers and solutions relevant to low resource settings.) *

**B**	Which of the following are the recognised key components for behaviour change strategy? (Select maximum 3) *

- Capability
- Motivation
- Opportunities
- Skills
- Knowledge
- I do not know
- Other: _________________________________________________________

**C**	Select which of the following are included in the ‘5 moments of hand hygiene’ according to WHO? (select all that apply)

- I do not know
- Before touching a patient
- before clean/aseptic procedures,
- after body fluid exposure/risk
- after touching a patient
- after touching patient surroundings
- after adjusting the IV rate and before checking blood pressure

**D**	What is the appropriate minimum length of time required for adequate hand hygiene

- I do not know
- 5 seconds for alcohol-based hand rub, 15 seconds for hand washing
- 15 seconds for hand washing, 5 seconds for alcohol-based hand rub
- 30-40 seconds for alcohol-based hand rub, 40-60 seconds for hand washing
- 60 seconds for alcohol-based hand rub, 60 seconds for hand washing
- None

**Appendix II**

**HEALTHCARE PROVIDERS’ INTERVIEW TOPIC GUIDE**

**Date of interview:**

**Respondent ID:**

**Sex:**

**Age range:**

**Specialty:**

**Length of time in Practice:**

**Introduction (to be read by interviewer just before commencing the interview):**

Thank you for accepting to take part in this interview. I would just like to reiterate that everything you say in the interview is confidential. All data collected will be anonymized. The interview itself will be open ended and the questions themselves are usually fairly broad. There aren’t any right or wrong answers. I’m simply interested in your experience and your views with antibiotics use. Is there anything you’d like to ask me?

**Did you attend the training session on AMR/Stewardship in July 2019? Y/N (if no skip next section)**

**Knowledge/Awareness of antimicrobial resistance post training**

1) Generally, how was this training helpful to you?
2) How did the training impact on your knowledge of AMR?
3) Has your practice with antibiotics changed since the training?
4) What are specific changes, what do you do differently?
5) Can you tell me about the WHO AWaRE classification of antibiotics?

[***List of Access, Watch and Reserve categories]***

6) We had a Global point prevalence survey (GPPS) on antibiotics in July, how has this affected your practice and understanding of antibiotics use?

**Pharmacist’s roles:**

1) In your opinion, how do pharmacists fit in AMS in hospitals?
2) Do you feel competent to deliver AMS roles within the hospital as a pharmacist/nurse/doctor/laboratory scientist? **[*Explore reasons].***
3) Are there enough resources within the hospital to help pharmacists be more efficient in doing this?
4) What would you recommend [resources/ structures] to support hospital pharmacists on AMS delivery?

**Knowledge/Awareness on antibiotics use in Ghana**

1) What is your general opinion on antibiotics use in Ghanaian hospitals?
2) Can you explain to me some of the factors that contribute to the current practices? [availability and access to antibiotics; lack of regulation; OTC availability; diagnostics; general knowledge expectations; cost; conflict of interest –business model]
3) To your knowledge are there any interventions put in place or any steps taken to help with antibiotics use in this hospital?

Probe: Any form of stewardship programmes? Antibiotic prescribing policies/ guidelines? Infection control groups; patient and public information campaigns; staff knowledge and awareness.

**Prescribing patterns**

1) Tell me about the antibiotics that are commonly used in this hospital, what are the common classes prescribed?
2) Are the prescribed antibiotics in the essential medicines list and hospital formulary?
3) What are the common indications requiring antibiotic prescriptions?
4) In your hospital practice, do you normally use antibiotics by brand names or by generic names…tell me about any reasons or factors that will affect generic or brand name use?

**Decision making process**

1) What are the factors that influence the choice of antibiotics you prescribed and dispensed? (Probes: Tests results, PC symptoms diagnosis, availability, cost, co- morbidities).
   a) What factors do you consider relevant in choosing antibiotics?
2) Are there treatment guidelines or antibiotic policies available to guide antibiotics use?
3) How often do you consult guidelines/policies/official books when dealing with antibiotics in the hospital?
4) In your opinion, do patients have any role to play in deciding if/ which antibiotic should be prescribed and dispensed to them?

**Recommended strategies**

1) In your opinion what interventions should be implemented in this setting to improve antibiotics use? Toolkits or checklists as potential facilitators?
2) What would be the likely barriers in implementing this intervention(s)

**APPENDIX III: AMS Interview data analysis**

| **S/N** | **Question** | **Responses/Codes** | **Key messages /Themes** |
| --- | --- | --- | --- |
| **Knowledge/Awareness of antimicrobial resistance post training** | | | |
| 1 | Generally, how was this training helpful to you? | Increased my ABX use knowledge  Very educative  Yes, very helpful  Yes, I learnt a lot, it was very very helpful.  very effective and helpful | Hcps described the training as educative, helpful and found it useful in their practice with antibiotics |
| 2 | How did the training impact on your knowledge of AMR? | Increases understanding of factors impacting AMR  Importance of patient counselling on abx use  I can now educate my patients better  It created another opportunity for us to help us with how antimicrobials will be used  it came to increase the knowledge that I had in AMR   impacted my knowledge of AMS yes  I have better understanding on the use of abx | The training generally increased knowledge about antibiotics use, AMR and AMS. Led to better understanding of different aspects of antibiotics use including importance of proper patient education |
| 3 | Has your practice with antibiotics changed since the training? | Personally yes  -Reduced antibiotics use,  - yes, I do more counselling with Abx drugs  Yes-reduced empirical abx prescribing  I should say yes,  Yes, I educate patients better now | After the training, some changes in practice observed in the hospital include reduction in antibiotics use and empirical prescribing.  Better patient counselling and education on antibiotics use |
| 4 | What are specific changes, what do you do differently? | Complete my course of ABX and advice others too  Abx prescription checks  I educate patients more on Abx, I contribute during ward rounds and query some abx prescriptions  We now query some antibiotic prescriptions without justification  Prescription checks for antibiotics  I stopped telling people to take antibiotics casually  actually, decreased the rate at which they were writing antibiotics | HCPs feel more confident to contribute on AMR topics during rounds and educate patients better. Pharmacists carry out prescription checks for antibiotics prescribed and query antibiotics prescribed where there is a need too |
| 5 | Can you tell me about the WHO AWaRE classification of antibiotics?  ***List of Access, Watch and Reserve categories*** | No, No, No, yes the category, Yes I remember the idea of this in 3 groups  Yes I remember 3 groups,  Yes Aware,watch and one other group | Majority of the HCP that attended the earlier training vaguely remembered the WHO classification of antibiotics |
| 6 | We had a Global point prevalence survey (GPPS) on antibiotics in July, how has this affected your practice and understanding of antibiotics use? | Yes, improved prescribing  Yes, evidence-based prescribing  Reduced broad spectrum empirical prescribing  Prescribers are more aware  Pharmacists provide input in abx prescribing confidently now  It has been difficult | The GPPS conducted made prescribers more aware of their prescribing with regards to antibiotics. There was reduce empirical and broad-spectrum antibiotic prescribing |
| **Pharmacist’s roles:** | | | |
| 7 | In your opinion, how do pharmacists fit in AMS in hospitals? | Nurse, yes, proper administration,  Support other HCP with information e.g prescribers  Pharmacists: drug custodians, ensure safety/storage and patient counselling  Yes, we procure the drugs and can influence the antibiotics in hospital | Pharmacists are the custodians of drugs within the hospital and can ensure safe effective use of antibiotics by providing information to other HCPs, counselling patients |
| 8 | Do you feel competent to deliver AMS roles within the hospital as a pharmacist/nurse/doctor/laboratory scientist? **[*Explore reasons].*** | Nurse yes- I administer ABX, so I can educate patients about it  Yes. With the right resources and support  As a nurse-support prescriber especially the junior doctors  Nurse- can act as patients advocate and question antibiotics being prescribed  Yes. I’m very very confident as a pharmacist now | Nurses are competent in abx drug administration and act as patient advocate to question antibiotics being prescribed  Pharmacists are also competent to deliver AMS roles and provide medication information needed to rational Abx prescribing |
| 9 | Are there enough resources within the hospital to help pharmacists be more efficient in doing this? | Yes, guidelines and other HCP  Labs should be equipped to deliver test results on time  The resources are not there to carry out the testing and all that in the lab,  Also, staff shortage: the pharmacist too, we are not many. | While there are guidelines and consultation with other colleagues, HCPs confirmed other resources like improved laboratory services, and increase in manpower will increase efficiency in delivery of AMS roles |
| 10 | What would you recommend [resources/ structures] to support hospital pharmacists on AMS delivery? | ABX use protocols/ guidelines  Training/ lab/diagnostics  Continuous training like CPD/workshops  Leaflets for patients, flip charts for HCP  AMR refresher courses  the disc for the culture and sensitivity, | HCPs recommended protocols and guidelines for ABX use, more AMR training courses, improved lab services and availability of patient educational materials to support the delivery of AMS in the local hospital |
| **Knowledge/Awareness on antibiotics use in Ghana** | | | |
| 11 | What is your general opinion on antibiotics use in Ghanaian hospitals? | Over usage, improvements in Abx use  Broad spectrum, empirical prescribing  Still need to work on use, reach people  Irrational use, poor IPC practices  There is a lot of antibiotic overuse, antibiotics are overused | HCPs were of the perceptions that there is a general there is irrational and overuse of antibiotics in Ghanaian hospitals citing broad spectrum and empirical prescribing of antibiotics |
| 12 | Can you explain to me some of the factors that contribute to the current practices? | Poor compliance to guidelines, patient demands, incentives from drug companies  Lack of training for HCPs, limited diagnostics  Poor knowledge on AMR, staff are overworked and have no time to consult or check before prescribing abx | Factors contributing to irrational use of antibiotics in Ghana include lack of continuous AMR trainings, poor AMR knowledge, limited diagnostic services, understaffing, pressure from patients and pharmaceutical companies |
| 13 | To your knowledge are there any interventions put in place or any steps taken to help with antibiotics use in this hospital? | NO, None (only some workshops) | There has been no specific interventions implemented to combat AMR in the hospitals |
| **Prescribing patterns** | | | |
| 14 | Tell me about the antibiotics that are commonly used in this hospital, what are the common classes prescribed? | Cephalosporins, penicillins, metronidazole, ciprofloxacin, erythromycin, amoxiclav, cefazolin, cipro | Broad-spectrum antibiotics including penicillin’s, cephalosporins aminoglycosides and metronidazole are commonly prescribed |
| 15 | Are the prescribed antibiotics in the essential medicines list and hospital formulary? | Yes, yes, yes, yes | Antibiotics prescribed in the hospital are in the EML and hospital formulary |
| 16 | What are the common indications requiring antibiotic prescriptions? | UTI, resp. TI, GIT infections, prophylaxis (after surgery), sepsis, STI, typhoid | Common indications for which antibiotics are prescribed include respiratory tract infections, UTI, STI, GIT infections. Others include typhoid and surgical prophylaxis |
| 17 | In your hospital practice, do you normally use antibiotics by brand names or by generic names…tell me about any reasons or factors that will affect generic or brand name use? | Both, generic mostly, both, generic, more of generic, both | Antibiotics are prescribed both as the brands and in generics but mostly in generics |
| **Decision making process** | | | |
| 18 | What are the factors that influence the choice of antibiotics you prescribed and dispensed? | Disease, cost, availability, severity of infection | The following factors are considered in making choice of antibiotics for a patient: the disease and its severity, cost of the antibiotic and availability. |
| 19 | Are there treatment guidelines or antibiotic policies available to guide antibiotics use? | Yes, we have our standard treatment guidelines | Presently, there are standard treatment guidelines in the hospital to guide antibiotics use |
| 20 | How often do you consult guidelines/policies/official books when dealing with antibiotics in the hospital? | As often as I need to  Yes, I do a lot, I now use the micro-guide app  As often as I need to | Most HCP employ the use of guidelines as often as they need to when dispensing, prescribing or administering antibiotics |
| 21 | In your opinion, do patients have any role to play in deciding if/ which antibiotic should be prescribed and dispensed to them? | No patients don’t have a say  No influence from patients  Yes, patients do request antibiotics, in the community they buy it | While patients can request for antibiotics, they have no influence in the decision whether they eventually get an antibiotic or which antibiotic should be prescribed for and dispensed to them. |
| **Recommended strategies** | | | |
| 22 | In your opinion what interventions should be implemented in this setting to improve antibiotics use? Toolkits or checklists as potential facilitators? | EDUCATION, more public awareness, policies  Checks/audits  Improved labs for microbe specific treatment, purchase lab items for testing  Training for prescribers  Monitoring antibiotic use  Employ more staff to build workforce,  Upgrade laboratory services | This will be presented and discussed using the COM-B model of the BCW framework for behaviour change |
| 23 | What would be the likely barriers in implementing this intervention(s) | Staff shortages, failure to enforce laws, funding  Staff Workload, lack of funding in healthcare | Same as above |
| Notes | | | |

**Appendix IV: Sample transcript Coding frame**

| **Coding frame for (Transcript T011)** | | |
| --- | --- | --- |
| **Questions** | **Quotes** | **Codes** |
| **Knowledge/Awareness of antimicrobial resistance post training** | | |
| Generally, how was this training helpful to you? | *“It was very very helpful”*  *“it came to create another opportunity for us to help us with how antimicrobials will be used”* | Useful, helpful |
| How did the training impact on your knowledge of AMR? | *“Well, initially I had some knowledge about AMR but it came to increase the knowledge that I had in AMR, it boosted the knowledge that was very very high and I think it was a very good programme”* | Increased AMR knowledge |
| Has your practice with antibiotics changed since the training? | *‘My practice has changed but it’s very difficult, especially in the hospital*”  *“some issues are still not being able to be solved, so there are no... I mean, the lab is not adequately resourced to test for the organisms and then for you to know the antibiotics to give and all that, so there are still some lapses”* | Yes, limited by other factors |
| What are specific changes, what do you do differently? | *“Yeah. But I mean, it does actually decrease the rate at which they were writing antibiotics”* | Reduction on antibiotic prescribing |
| Can you tell me about the WHO AWaRE classification of antibiotics?  [***List of Access, Watch and Reserve categories]*** | *“Yes. The WHO AWaRe. So they have the Access, the Watch and then...I’m trying to remember”* | Partially |
| We had a Global point prevalence survey (GPPS) on antibiotics in July, how has this affected your practice and understanding of antibiotics use? | *“Yes. Yes-yes-yes-yes, it has certainly helped .. In lots of ways. So now, I mean, the prescribers are aware and we also give them... I mean, if they need our input, we also give them our input on which antimicrobials are to be used for some conditions, so it has really...”* | Increase prescriber awareness |
| **Pharmacist’s roles:** | | |
| In your opinion, how do pharmacists fit in AMS in hospitals? | *“Well the pharmacist has a critical role to play because we are the custodians of drugs”*  *“can actually select some drugs, appropriate medications to be procured by the hospital so that they can factor in they want to prescribe an antibiotic and we can still be educating them, the prescribers, on right use of the antimicrobials, so as to prevent any resistant strains from developing”* | Support prescribers  Procure appropriate antibiotics |
| Do you feel competent to deliver AMS roles within the hospital as a pharmacist/nurse/doctor/laboratory scientist? **[*Explore reasons].*** | *“Yes. I’m very very confident.”* | Yes, confident |
| Are there enough resources within the hospital to help pharmacists be more efficient in doing this? | *“Like I said, the resources are not there, you go to the lab, there are no agents, they don’t have a disc to carry out the testing and all that,*  *“The pharmacist too, we are not many”* | Limited lab resources  Staff shortage |
| What would you recommend [resources/ structures] to support hospital pharmacists on AMS delivery? | “*We need all... I mean the disc for the culture and sensitivity, very very important because if you are advocating that they change from one antibiotic to the other and there is no evidence*  *Now we are going by the evidence, so we need the culture and sensitivity testing to help you, so it’s very very... I think”* | Improved lab resources |
| **Knowledge/Awareness on antibiotics use in Ghana** | | |
| What is your general opinion on antibiotics use in Ghanaian hospitals? | *“it’s a major problem here in Ghana, a big issue here in Ghana because when you go to the hospital, I mean, almost every patient, at least almost every patient we put on an antibiotic”*  *“Yes. There is a lot of antibiotic overuse and the sad aspect with that, they don’t start from... I mean, you know the Accessible, the Watch and then the Reserve”* | Antibiotic overuse |
| Can you explain to me some of the factors that contribute to the current practices? | *“as I mentioned earlier, resources are not available”*  *“you have a lot of patients waiting for you, so you just go like working”* | Limited lab resources  Staff shortages |
| To your knowledge are there any interventions put in place or any steps taken to help with antibiotics use in this hospital? | *“I think more resources should be available to help so that the lab scientists”*  *“Then more pharmacists should also be employed”* |  |
| **Prescribing patterns** | | |
| Tell me about the antibiotics that are commonly used in this hospital, what are the common classes prescribed? | *“So Amoxiclav and then Cefazolin*  *Those two, almost every patient in Ghana who has been admitted at the hospital has been... They’ve been on that medication.”* | Broad spectrum antibiotics |
| Are the prescribed antibiotics in the essential medicines list and hospital formulary? | *“Yeah-yeah, they are.”* | Yes, on the EML |
| What are the common indications requiring antibiotic prescriptions? ***{Review the need for this}*** | *“Upper respiratory tract infection and STI”* | URTI and STIs |
| In your hospital practice, do you normally use antibiotics by brand names or by generic names…tell me about any reasons or factors that will affect generic or brand name use? | *“We use both. We use both brand name and generic”.* | Both |
| **Decision making process** | | |
| What are the factors that influence the choice of antibiotics you prescribed and dispensed? | *“So the issue is that I have my colleagues, the other medical doctors who comes to you and say that they think the generic is not working for a patient, they think it doesn’t contain the right amount of... I mean the right strength in the medication because they keep giving it to the person but it doesn’t work. But the moment they switch from let’s say the generic to a brand, then they begin to get some...”* | Perceive brands are more effective |
| Are there treatment guidelines or antibiotic policies available to guide antibiotics use? | *“Yes. So from our standard treatment guidelines, for every infection they have their first line that are to be used, they have second line there, sometimes third line they have for that infection, so we try as much as possible to follow.”* | STG available |
| How often do you consult guidelines/policies/official books when dealing with antibiotics in the hospital? | *“For me every time I’m at the ward I check.*  *I make sure that what you are doing, I mean if they’re able to give me the right diagnosis that this infection is let’s say pneumonia, bronchial pneumonia, then I’ll be able to tell the right antibiotic to be used.”* | As often as needed |
| In your opinion, do patients have any role to play in deciding if/ which antibiotic should be prescribed and dispensed to them? | *“Well sometimes, yes, a patient can actually push for that”*  *“…. they just want an antibiotic. Sometimes they’ll push you to prescribe an antibiotic”* | Patient pressure |
| **Recommended strategies** | | |
| In your opinion what interventions should be implemented in this setting to improve antibiotics use? Toolkits or checklists as potential facilitators? | *“I’ve answered this in a different way but I think the first one should be the resources, the resources should be available, human resources”*  *“I mean, for them to be able to go by, so that they will be able to do a culture and sensitivity test, so that they’ll be able to go for the right antibiotic to be used”*  *‘then as a pharmacist, if I’m doing my... In my role I’ll be able to guide them appropriately”* | Improved labs  Manpower |
| What would be the likely barriers in implementing this intervention(s) | *“Obviously, I mean money will play a key issue. Yes, so our finances”* | Funding |
